# Integrated MB-PhD training is a long-term investment in the clinician-scientist workforce

**DOI:** 10.64898/2026.08.26.26361003

**Authors:** Daniyal J Jafree, Mingze Sun, Gordon W Stewart, Faye Gishen, Charles Swanton, Reza Motallebzadeh, the UCL MB-PhD Outcomes Study Group

**Affiliations:** Development Biology & Cancer Research and Teaching Department, UCL Great Ormond Street Institute of Child Health, London, UK; UCL Centre for Kidney & Bladder Health, University College London, London, UK; Department of Women’s and Children’s Health, University of Liverpool, Liverpool, UK; Wellcome Sanger Institute, Wellcome Genome Campus, Hinxton, UK; The Hatter Cardiovascular Institute, University College London, London, England, UK; Division of Medicine, University College London, London, UK; UCL Medical School, University College London, London, UK; Whittington Health NHS Trust; Cancer Evolution and Genome Instability Laboratory, The Francis Crick Institute, London, UK; Cancer Research UK Lung Cancer Centre of Excellence, University College London Cancer Institute, London, UK; Research Department of Surgical Biotechnology, University College London, London, UK; Institute of Immunity and Transplantation, University College London, London, UK; Department of Nephrology and Transplantation, Royal Free Hospital, London, UK

**Author notes:** Joint corresponding authors **Dr Daniyal J Jafree**, Senior Research Fellow, UCL Great Ormond Street Institute of Child Health, London, UK, NIHR Academic Clinical Fellow, University of Liverpool, Liverpool, UK, Senior Research Fellow, Wellcome Sanger Institute, Hinxton, UK, **Professor Reza Motallebzadeh**, Professor of Renal Transplantation, Department of Surgical Biotechnology, UCL, London, UK, MB-PhD Programme Director, University College London, London, UK, Honorary Consultant Renal Transplant Surgeon Royal Free Hospital, London, UK.

## Abstract

**Background:** Clinician-scientists translate clinical observation into discovery, trials, and policy, yet this workforce is shrinking across health systems worldwide. Integrated MB-PhD training, pausing medical training to complete a PhD before clinical exposure or specialisation, is one route into this career. We aimed to evaluate the long-term value of MB-PhD training and the barriers to clinical-academic careers faced after graduation.

**Methods:** We evaluated all 131 graduates (29.8% female) who entered the University College London (UCL) MB-PhD programme over a 25-year period (1994-2018). Bibliometric outputs were collated via an inter-linked information system. Concurrently, all 131 graduates were invited to respond to open-ended questions on career benefits and structural barriers; 99 (75.6%) responded, and responses were independently coded into themes, which were then reviewed and confirmed by a Study Group of 107 individuals, including the 91 respondents who agreed to participate further.

**Results:** Graduates produced 5,877 publications (1,141 first-author, 819 corresponding-author), attracting 350,754 citations, with a mean relative citation ratio of 3.30 ± 0.47, approximately three times the field average and sustained across three decades of programme entry. Graduates secured an estimated $157.55 million across 99 grants, released 465 public datasets, and were named investigators on 31 clinical trials across five continents. Among the 99 survey respondents, 49.5% held consultant-grade posts, 72.7% remained research-active, and 25.3% had reached senior academic grade. Openended responses were coded into five recurring structural barriers, subsequently confirmed by the Study Group: insufficient protected research time (72.2% of responses), unsupportive training structures and limited career opportunities (36.7%, 24.4% of responses), funding and pay barriers (22.2% of responses), and lack of mentorship or geographical/family constraints (14.4%, 13.3% of responses).

**Conclusions:** Integrated MB-PhD training generates sustained academic productivity and leadership, but structural barriers threaten retention of graduates within clinical-academic careers. Protecting research time, stabilising funding and pay, and reducing geographic instability, amongst other interventions, are needed to retain the clinician-scientists that health systems have already invested in training.

---

Clinician-scientists translate clinical observations into discovery, trials and policy towards patient benefit. However, this workforce is shrinking across health systems, adversely impacting the health of nations^1^. In the UK, the number of clinical academics under 36-years-old fell by 25% from 2024-2025^2^, threatening the pipeline needed to sustain research capacity as senior academics retire^3^. Accordingly, the proportion of clinical academic consultants declined, from 4.7% in 2009 to 3.2% in 2025^2^. Similarly, attrition of clinician-scientists is documented in the USA, Europe and Australia, particularly at career transitions^4-6^. Globally, health systems are failing to retain those trained to deliver translational medicine and the expertise bridging academic discovery with industry-led drug development to improve patient care.

One of several routes to becoming a clinician-scientist involves pausing early medical training to complete a PhD in discovery research. Referred to in the UK as MB-PhD training, these programmes offer deep research experience before clinical exposure or specialisation. We aimed to examine what early commitment to clinician-scientist training, such as MB-PhD programmes, can achieve and the barriers to clinical-academic careers these graduates face.

Using an inter-linked information system to collate bibliometric outputs, we evaluated all 131 graduates (29.8% female) who entered MB-PhD training over a 25-year period (1994 to 2018 inclusive) at University College London (UCL)^7^, the UK’s second oldest MB-PhD programme after Cambridge^8^. These 131 individuals produced 5,877 publications (4,851 original articles, 714 preprints, 155 conference proceedings, 154 book chapters), including 1,141 firstauthor and 819 corresponding-author papers, attracting 350,754 citations. The mean relative citation ratio (RCR) was 3.30 ± 0.47, approximately three times higher than work in related fields, and sustained across graduates grouped by time period from programme entry (1994-2001 entry: 2.67 ± 0.49; 2002-2009 entry: 3.89 ± 1.09; 2010-2018 entry: 3.36 ± 0.67, **Fig.1a**, distributional statistics in *Appendix*). Graduates secured ∼ $157.55 million across 99 grants, likely underestimated, since not all grants are documented publicly, released 465 public datasets, and were named investigators on 31 clinical trials registered across five continents.

**Figure 1:**
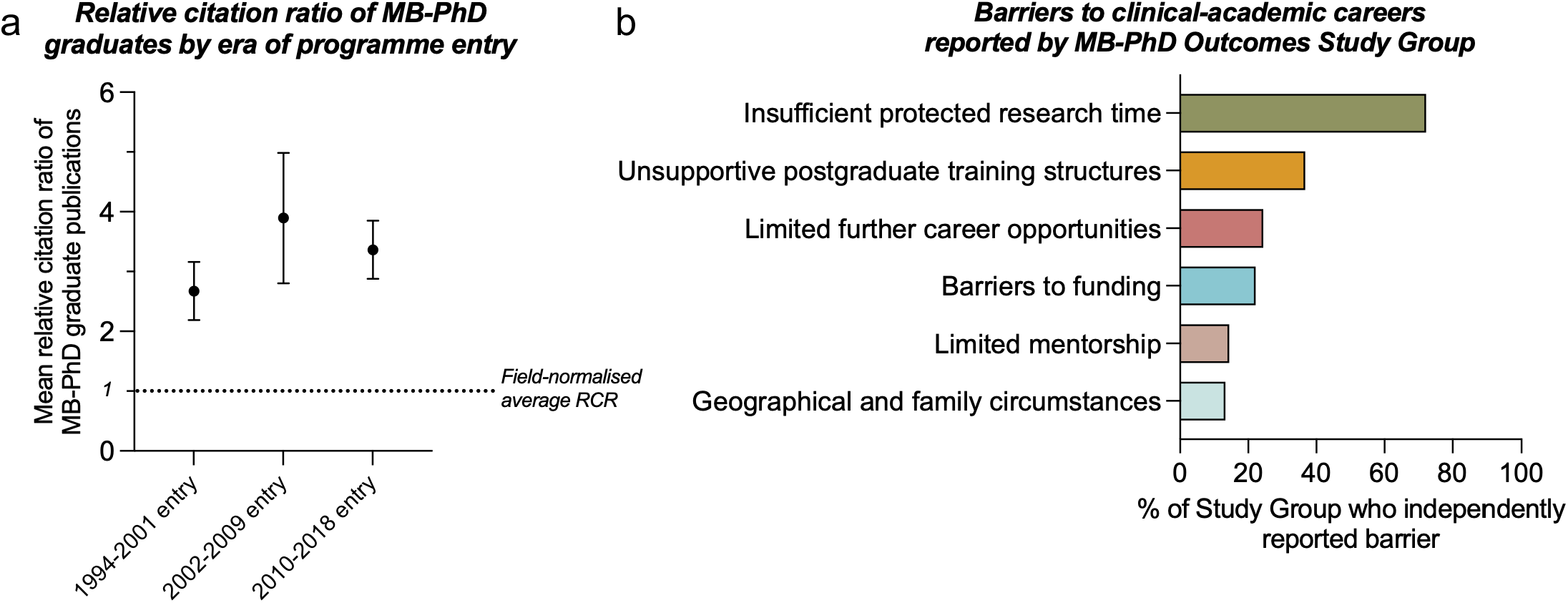
Long-term research impact of MB-PhD graduates and barriers to clinical academic careers. (**a**) Mean relative citation ratio (RCR) among graduates, stratified by year of entry to the MBPhD programme: 1994-2001 entry: 2.67 ± 0.49; 2002-2009 entry: 3.89 ± 1.09; 2010-2018 entry: 3.36 ± 0.67. The dotted line indicates the field-normalised reference value of 1. Points represent mean RCR and error bars indicate the standard error of the mean. (**b**) Five most frequently reported consensus barriers to pursuing a clinical academic career, through an open question format. Values indicate the proportion of respondents whose responses were assigned to each barrier. Individual responses could be assigned to more than one barrier.

Concurrently, we invited all 131 graduates to participate in an evaluation of integrated MB-PhD training. Of 131 graduates, 99 (75.6%) responded, with demographics consistent with Cambridge MB-PhD graduates^9^: 49.5% held a consultant-grade post, 72.7% remained research-active, and 25.3% had reached Professor, Associate Professor, or Senior Lecturer level, of whom 24.0% were women. A further 12.1% held clinical lectureships, of whom 33.3% were women. 60.2% entered a clinical specialty different from the closest match to their PhD, challenging the assumption that early doctoral training only matters if graduates remain in the same research field. Instead, graduates reported that integrated MB-PhD training strengthened career progression, transferable skills and access to alternative careers, consistent with US cohorts^10^.

Participating graduates identified structural barriers (**Fig.1b**) to clinical-academic careers, reviewed and refined by our MB-PhD Study Group comprising 91/99 respondents, with responses grouped into five recurring challenges echoing reports by the Office for the Strategic Coordination of Health Research (OSCHR)^11^ and European^4^ and Australian^6,12^ MD-PhD cohorts.

First, 72.2% cited insufficient protected research time and overwhelming clinical workload preventing research progression. A system that supports early research training but does not allow protected academic time after graduation will inevitably interrupt productivity, compounding attrition at the key career transitions already documented internationally^4^.

Second, 36.7% cited unsupportive training structures and 24.4% cited limited career opportunities, including pathways designed without reference to prior doctoral training. Formal pathways in the UK, Germany and France^12^ enable protected research time, but do not always accommodate graduates who already hold substantial prior research experience. Our Study Group’s clinical-academics report that a clinical lectureship is often unavailable until 5-10 years after PhD completion, forcing graduates to rebuild a research career from an earlier stage than their training justifies. This disconnect is exemplified by the Specialised Foundation Programme (SFP) in England, where two-thirds of posts are now allocated randomly rather than by merit^13^, meaning MB-PhD graduates who have already demonstrated research productivity may be selected no differently to those without any.

Third, 22.2% cited funding barriers. Study Group members reported scarce protected salary support between PhD completion and independent fellowship acquisition, and no clear route onward at the end of the training pathway, given low intermediate fellowship success rates and inconsistent research time within consultant job plans. Inadequate pay structures were also highlighted, which penalise doctoral training completed before, rather than during, specialty training. The 2016 Doctors’ and Dentists’ Terms and Conditions determine eligibility for an academic pay premium^14^ from which many Study Group graduates report exclusion, a problem compounded by global funding instability.

Fourth, 14.4% cited lack of mentorship and 13.3% cited geographical and family circumstances. Widening geographical distribution of clinical-academic posts may address regional inequalities, but the resulting instability is detrimental to graduates with established mentorship and research networks. This is particularly challenging for those with domestic caring responsibilities. Gender disparities in MD-PhD career progression are well documented^15^ and how geographic instability drives this warrants further attention.

Our analysis has caveats. Senior academics may be over-represented in bibliometric analyses, typical of citation data, though median RCR exceeded the field average in every era of programme entry. All graduates originated from a single MB-PhD programme without a comparator group. Furthermore, we cannot exclude that motivated individuals would have succeeded regardless of programme participation. Additionally, ethnicity data was not recorded and Study Group participants may be more academically engaged than non-participants.

Taken together with experiences from the UK^9^, Europe^4^ and North America^6,10^, our findings suggest that integrated MB-PhD training generates sustained academic productivity and leadership. Realising the value of clinician-scientist programmes, including MB-PhD training, and preventing loss of essential translational research expertise for healthcare and pharmaceutical drug development, depends on whether action is taken to retain those already trained. This requires protected research time, sustained funding, flexibility in balancing research with specialty training, pay that recognises research experience regardless of when it occurs and posts that do not force graduates to distance from their research. Coordination between governments, healthcare systems, funders and universities should ensure that MBPhD graduates, and clinician-scientists more widely, shape workforce policies that govern their careers.

## Data Availability

Bibliometric data are indexed in publicly accessible databases, though aggregated analysis required licensed access via the Dimensions platform. Aggregated and coded analyses of responses to open-ended questions on career benefits and structural barriers of integrated MB-PhD training are available upon reasonable request to the authors.

## Competing Interests and Acknowledgements

DJJ, GWS, FG, CS and RM designed the study, supported by members of the MB-PhD Outcomes Study Group. DJJ, MS and GWS performed data collection. DJJ, MS and RM analysed results. DJJ and RM wrote the first draft of the paper, and subsequently, all co-authors, including the MB-PhD Outcomes Study Group, revised and approved the final version of the manuscript. DJJ, MS and CS are UCL MB-PhD programme alumni. RM is current Director; GS is former and founding Director and FG is Director of UCL Medical School. The authors declare no other competing interests relevant to this article. The bibliometric data in this manuscript were sourced from Dimensions, an interlinked research information system provided by Digital Science (https://www.dimensions.ai) under their Scientometric Researcher Access to Data (SRAD) programme (data extracted on 4^th^ July 2026). We acknowledge Professors Tony Segal, Neville Wolff and David Brenton, who founded the UCL MB-PhD programme and facilitated its integration into UCL Medical School, along with Professors David Katz, Patrick Maxwell, Mary Collins, Jane Dacre, Geraint Rees, Emma Morris, Patrick Vallance and Ms Gaynor Jones. We also thank Professors Stefan Marciniak and Timothy Cox (University of Cambridge MB-PhD programme) for invaluable discussions. Investment in MB-PhD studentships, the programme or its students and graduates has been received from the Francis Crick Institute, the Wellcome Trust, Cancer Research UK, the British Heart Foundation, the LifeArc Centres for Rare Diseases, Kidney Research UK, the Kennedy Trust, the Rosetrees Trust, the International Journal of Experimental Pathology, the Astor Foundation, the Jean Shanks Fund, A*Star (Singapore), the Sir Jules Thorn Trust, the Lord Amulree Fund, the Foulkes Foundation, UCLH Charity and GSK.

