## Appendix for "Integrated MB-PhD training is a long-term investment in the clinician-scientist workforce"

[+] Joint corresponding authors

**Dr Daniyal J Jafree**

Senior Research Fellow, UCL Great Ormond Street Institute of Child Health, London, UK

NIHR Academic Clinical Fellow, University of Liverpool, Liverpool, UK

Senior Research Fellow, Wellcome Sanger Institute, Hinxton, UK

**Professor Reza Motallebzadeh**

Professor of Renal Transplantation, Department of Surgical Biotechnology, UCL, London, UK

MB-PhD Programme Director, University College London, London, UK

Honorary Consultant Renal Transplant Surgeon Royal Free Hospital, London, UK

**Table of contents**

***MB-PhD Outcomes Study Group co-authors and affiliations*** **3**

***Methods*** **12**

***Supplementary data*** **14**

***References*** **16**

**MB-PhD Outcomes Study Group co-authors and affiliations**

**Addenbrooke's Hospital, Cambridge, UK**

Richard Mannion

**Albert Einstein College of Medicine, New York, USA**

Joshua Kahan

**AstraZeneca, UK**

Daniel JB Marks

**Barts Health NHS Trust, London, UK**

Corrine Stannard

**Bioheng Biotech, Nanjing, China**

Jan Davidson-Moncada

**Blackrock Health Blackrock Clinic, Dublin, Ireland**

Shamindra Direkze

**Cancer Research UK Centre for Drug Development, London, UK**

Alvin J X Lee

**Central Health London, London, UK**

Benjamin Davis

**Chelsea and Westminster Hospital NHS Foundation Trust, London, UK**

Alejandro Acosta-Saltos

Andrew Godfrey

**Clinique Saint-Jean, Brussels, Belgium**

Eugenia Papakrivopoulou

**Dubai Health, Dubai, United Arab Emirates**

Nadia Taysir Dabbagh

**GSK, London, UK**

Daniel Swerdlow

**Haga Hospital, The Hague, Netherlands**

Marieke Bokhoven

**Harley Street Maxillofacial Clinic, London, UK**

Kaveh Shakib

**Hebrew University-Hadassah Medical Center, Jerusalem, Israel**

Adam P Levine

**Hunter New England Local Health District, New South Wales, Australia**

Tom MacDougall

**Imperial College London, London, UK**

Elisabeth Robinson

Marcus Ghosh

Stephen D Auger

Vishal Rawji

**Institute of Mental Health, Singapore**
Geoffrey Chern-Yee Tan

**King's College Hospital NHS Foundation Trust, London, UK**
Ryan Wee

**King's College London, London, UK**

Dhruva Biswas

Isobel Weinberg

Joel S Winston

Matthew Wright

Michael Lim

Rafael Di Marco Barros

Sean E Cavanagh

Yin Wu

**London North West University Healthcare NHS Trust, London, UK**
Panagiotis G Kyrtatos

**Luton and Dunstable University Hospital, Luton, UK**

Chrysostomos Tornari

**Manchester Centre for Clinical Neurosciences, Salford, UK**

Rachel Thomasson

**Medicines and Healthcare products Regulatory Agency, London, UK**

King-Yin Lee

**Moorfields Eye Hospital NHS Foundation Trust, London, UK**

James RC Miller

**National University Hospital, Singapore**

Shao Jin Ong

**Newcastle upon Tyne Hospitals NHS Foundation Trust, Newcastle upon Tyne, UK**

Ina Schim Van Der Loeff

Lawrence Best

Luke La Hausse De Lalouviere

**NHS Fife, Fife, UK**

Devesh J Dhasmana

**Nottingham University Hospitals NHS Trust, Nottingham, UK**

Alexander Rossdeutsch

Paul Candler

**Queen Mary University of London, London, UK**

Malvina Cunningham

Sam Ereira

**Roche, United States**

Antonia Kwan

**Royal Adelaide Hospital, Adelaide, Australia**

Marcus Wagstaff

**Royal Berkshire NHS Foundation Trust, Reading, UK**

Shawn David P Ellis

**Royal Free London NHS Foundation Trust, London, UK**

Aine Burns

Sorena Kiani-Alikhan

**Royal Papworth Hospital NHS Foundation Trust, Cambridge, UK**

William Jenner

**Stanford University, Stanford, USA**

Thomas BK Watkins

**The Francis Crick Institute, London, UK**

Daniel Snell

James Turner

Rupert Beale

**The Walton Centre NHS Foundation Trust, Liverpool, UK**

Saba Rose Raza-Knight

**Universidad Politécnica de Madrid, Madrid, Spain**

Bryan A Strange

**University College London (UCL), London, UK**

Abhishek Das

Adam Pennycuick

Alexander Brown

Carolyn Cohen

David Katz

Gabriel Pollara

Izabella Smolicz

Joshua Carmichael

Lucy Bell

Mahdad Noursadeghi

Nigel Field

Quentin J M Huys

Raymond MacAllister

Robert Unwin

Sam Janes

Sophie Adler

Stavros Loukogeorgakis

Susan Beesley

Tony Segal

Uma Mukherjee

**UCL Hospitals NHS Foundation Trust, London, UK**

Ashkon Syed-Safi

Christian Hasford

Marc George

Ricardo José

**University Hospital Cologne, Cologne, Germany**
Lukas Heukamp

**University Hospital Southampton NHS Foundation Trust, Southampton, UK**
Weeratunge M N Malalasekera

**University Hospitals Bristol and Weston NHS Foundation Trust, Bristol, UK**
Adam C Nunn

**University Hospitals of Liverpool Group, Liverpool, UK**
James Arrich

**University Hospitals Plymouth NHS Trust, Plymouth, UK**
Sharon Man

**University of Birmingham, Birmingham, UK**
Victoria Wykes

**University of Bristol, Bristol, UK**

Adam Smith-Collins

Catherine Hyams

Jonathan Tyrrell-Price

Panayiotis Maghsoudlou

**University of Cambridge, Cambridge, UK**

Gavin W Sewell

Kate Baker

Mina Ryten

Oliver Patrick Devine

Rebecca A Burrell

Richard Bartlett

**University of Dundee, Dundee, UK**

Tom Gilbertson

**University of Liverpool, Liverpool, UK**

Lance Turtle

Reecha Sofat

**University of Manchester, Manchester, UK**

Sean Knight

**University of Oxford, Oxford, UK**

Anna Rose

Christopher Bricogne

Christopher McKinnon

Noura Al-Juffali

Thomas Parr

**Whittington Health NHS Trust, London, UK**

Catherine Storm

### Methods

#### *Bibliometric analysis*

Publication data for all 131 graduates who had successfully completed the MB-PhD, including completion of both PhD and MBBS components of the programme up until 2018 programme entry (to only include graduates who would have been expected in this timeline to complete the UK Foundation Programme), were extracted from Dimensions (Digital Science)[1], an interlinked research information system, under the Scientometric Researcher Access to Data (SRAD) programme (data extracted on 4th July 2026).

For programme-wide, aggregated metrics including number of publications, citation counts, grant funding and clinical trial involvement across all MB-PhD graduates, each graduate was manually searched and added to a ‘Group’ within the Dimensions browser.

For analysis of first and corresponding author publications, a spreadsheet of all MB-PhD co-authored publications was extracted, before matching first and corresponding author publication records using a custom Python script, identified by parsing the author list and corresponding-author fields of each publication record.

For relative citation ratio (RCR)[2], a mean RCR was ascertained for each graduate by Dimensions, and graduates were grouped into three time periods of programme entry (1994-2001, 2002-2009, 2010-2018) to generate longitudinal metrics on those that would be expected to have completed the UK Foundation Programme. Aggregated RCR values for each era of programme entry, and across the whole cohort of graduates, are presented as means ± standard error of the mean (SEM). Median values, kurtosis and skewness are shown in **Supplementary Data 1.**

#### *Determination of barriers to clinical-academic training*

The method to determine barriers to clinical-academic training is summarised by **Supplementary Data 2**. All 131 graduates were invited by email to participate in an evaluation of career outcomes and barriers associated with MB-PhD training, *via* a Microsoft Forms survey, using a combination of free-text and closed-question responses. The questions were as follows:

i) What year did you enter the MB-PhD programme?
ii) Did your PhD help you to achieve your career goals?

iii) What were your main challenges/barriers for continuing academic work during clinical training?
iv) What type of position do you currently hold? (Options: Clinical; Clinical & Research; Academia; Industry; Other)
v) Please select your current clinical grade
vi) What is your current clinical specialty?
vii) Please select your current academic grade
viii) What proportion of your current working time is dedicated to research?
ix) If you are engaged in research now, what type of research?

Ninety-nine of 131 graduates (75.6%) responded. Of these 99 respondents, 91 agreed to form the MB-PhD Outcomes Study Group. Answers to questions (ii) and (iii) were checked for anonymity, with names, positions and identifying information manually removed. ChatGPT (version 5.6, Luna) was used to assist with thematic analysis, by generating preliminary themes across identifiable information-redacted and anonymised responses. The proposed themes were then evaluated and manually edited by two independent researchers, before manual researcher-led assignment of each response to one or more of the themes. The compiled results were circulated to the remaining lead authors for review, before circulation to the Study Group as part of the manuscript draft. Feedback was sought from the Study Group and responses were refined before recirculation for final review. PhD thesis topic was manually assigned to the closest specialty by two researchers and compared to results from question (vi).

#### *Ethics statement and data acquisition*

Ethical approval was obtained through UCL's Life and Medical Sciences Research Ethics Committee (REC ID: 4751). Bibliometric data are indexed in publicly accessible databases, though aggregated analysis required licensed access via the Dimensions platform. Data from the survey and consensus process were provided voluntarily by MB-PhD graduates, all of whom had the opportunity to review, amend, or withdraw their contribution prior to submission.

#### *Statistics and data presentation*

Mean, SEM, median values and distributional analysis for RCR was calculated using Excel for Mac (Microsoft, Version 16.110.3). All graphs shown in the manuscript and Appendix were plotted using Prism for MacOS (GraphPad, Version 11.0.2).

### Supplementary data

| Era of entry | Mean RCR | Median RCR | SEM | Kurtosis | Skewness |
|---|---|---|---|---|---|
| **1994-2001** | 2.67 | 1.42 | 0.49 | 7.03 | 2.57 |
| **2002-2009** | 3.89 | 2.04 | 1.09 | 14.56 | 3.74 |
| **2010-2018** | 3.36 | 2.07 | 0.67 | 3.85 | 1.87 |
| ***Aggregated*** | *3.30* | *1.94* | *0.47* | 21.51 | 4.14 |

Supplementary data 1: Distributional metrics for relative citation ratio across MB-PhD graduates, grouped by era of entry and aggregated.

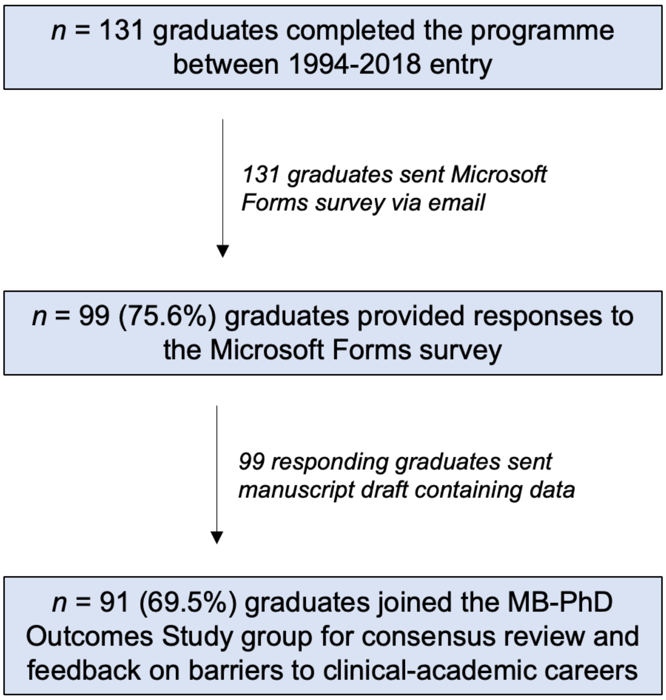


Supplementary data 2: Flow diagram demonstrating the method to survey and establish consensus amongst the MB-PhD Outcomes Study group.

| Journal | Number of publications |
|---|---|
| ***Nature*** | 63 |
| ***Cell*** | 28 |
| ***Science*** | 12 |
| ***The Lancet*** | 53 |
| ***New England Journal of Medicine*** | 14 |
| ***The British Medical Journal*** | 16 |
| ***Journal of the American Medical Association*** | 9 |

Supplementary data 3: Quantification of MB-PhD graduate co-authored publications in landmark discovery science and clinical research journals. Numbers represent programme-wide aggregates across all time-periods.

| Era of entry | Number (%) of males | Number (%) of females |
|---|---|---|
| **1994-2001** | 29 (70.7) | 12 (29.3) |
| **2002-2009** | 25 (61) | 16 (39) |
| **2010-2018** | 38 (77.6) | 11 (22.5) |
| **2019-2025** | 16 (47.06) | 18 (52.94) |

Supplementary data 4: Longitudinal uptake of male or female MB-PhD candidates over era of time of MB-PhD programme entry.

**References**

The two references used in this Appendix are cited below. In addition, we have provided a list of additional references that, due to space constraints, were not referenced in the manuscript, but are highly relevant to the topic of MB-PhD or MD-PhD programmes and the challenges clinical-academic trainees face in the current global healthcare landscape.

1. Bode C, Herzog C, Hook D, McGrath R, Wade A. A guide to the Dimensions data approach. Technical report. Digital Science; 2023. DOI: 10.6084/m9.figshare.5783094.
2. Hutchins BI, Yuan X, Anderson JM, Santangelo GM. Relative citation ratio (RCR): a new metric that uses citation rates to measure influence at the article level. PLoS Biol 2016; 14: e1002541.

***Additional references***

Academy of Medical Sciences. MB PhD programmes: a position paper by the Academy of Medical Sciences. London: Academy of Medical Sciences, 2007.

Akabas MH, Brass LF. The national MD-PhD program outcomes study: outcomes variation by sex, race, and ethnicity. JCI Insight 2019; 4: e133010.

Akabas MH, Brass LF. The National MD-PhD Program Outcomes Study: career paths followed by Black and Hispanic graduates. JCI Insight 2024; 9: e178248.

Akabas MH, Tartakovsky I, Brass LF. National MD-PhD Program Outcomes Study. Washington, DC: Association of American Medical Colleges, 2018.

Al Naimi A, Hentrich A, Bahlmann F, Louwen F. The impact of doctorate-timing, whether undergraduate or postgraduate, on the experience of German doctors. BMC Med Educ 2026; 26: 341.

Alamri Y. The combined medical/PhD degree: a global survey of physician-scientist training programmes. Clin Med (Lond) 2016; 16: 215–18.

Andriole DA, Whelan AJ, Jeffe DB. Characteristics and career intentions of the emerging MD/PhD workforce. JAMA 2008; 300: 1165–73.

Barnett-Vanes A, Ho G, Cox TM. Clinician-scientist MB/PhD training in the UK: a nationwide survey of medical school policy. BMJ Open 2015; 5: e009852.

Barreto-Duarte B, Mendelsohn SC, de Bruyn E, Andrade BB. What it takes to become a physician scientist in a low- and middle-income country. PLOS Glob Public Health 2025; 5: e0004234.

Batavanis M, Arora A, Aryee S, Nathwani N. Exploring barriers to expanding medical training numbers in England: a national survey of medical education directors. Clin Med (Lond) 2025; 25: 100523.

Brown OI, Morgan H, Jenner WJ, Chapman A, Joshi A, Drozd M, et al. Joint British Societies' position statement on cardiology training in the United Kingdom. Heart 2025; 111:e2.

Chakraverty D, Cavazos JE, Jeffe DB. Exploring reasons for MD-PhD trainees' experiences of impostor phenomenon. BMC Med Educ 2022; 22: 333.

Clough S, Fenton J, Harris-Joseph H, Rayton L, Magee C, Jones D et al. What impact has the NIHR Academic Clinical Fellowship (ACF) scheme had on clinical academic careers in England over the last 10 years? A retrospective study. BMJ Open 2017; 7: e015722.

Crispi V, Bolton W, Chand M, Giuliani S, Wykes V, Mathew RK. Barriers to clinical academic surgical training and career development in the United Kingdom: a review from the National Institute for Health and Care Research Advanced Surgical Technology Incubator Group. Br J Hosp Med (Lond) 2025; 86: 1–12.

Dos Santos Rocha A, Combescure C, Negro F. The MD-PhD program in Geneva: a 10-year analysis of graduate demographics and outcomes. BMC Med Educ 2020; 20: 447.

Eley DS. MD-PhD programs should encourage and support training in nonbiomedical or clinical sciences. Acad Med 2016; 91: 1038.

Eley DS, Jensen C, Thomas R, Benham H. What will it take? Pathways, time and funding: Australian medical students' perspective on clinician-scientist training. BMC Med Educ 2017; 17: 242.

Eley DS, Stormon N, Ford PJ. The clinician-scientist track: an approach addressing Australia's need for a pathway to train its future clinical academic workforce. BMC Med Educ 2018; 18: 227.

Forde A. An overview of German MD/PhD programs. Science Careers. Oct 10, 2003. https://www.science.org/content/article/overview-german-mdphd-programs (accessed July 21, 2026).

Ganem D. The MD/PhD: what comes after? Science Careers. Oct 24, 2003. https://www.science.org/content/article/mdphd-what-comes-after (accessed July 21, 2026).

Harding CV, Akabas MH, Andersen OS. History and outcomes of 50 years of physician-scientist training in Medical Scientist Training Programs. Acad Med 2017; 92: 1390–98.

Harvey Bluemel A, Burton OE, Burford B, Ellis J, Finn G, Bala L, et al. Barriers and facilitators to establishing a clinical academic career in clinical education research in the UK: a focus group study. Med Teach 2024; 46: 1369–77.

InterAcademy Partnership for Health, Academy of Medical Sciences. Strengthening clinical research capacity in low- and middle-income countries. London: Academy of Medical Sciences, 2017.

Jeffe DB, Andriole DA. A national cohort study of MD-PhD graduates of medical schools with and without funding from the National Institute of General Medical Sciences' Medical Scientist Training Program. Acad Med 2011; 86: 953–61.

Jeffe DB, Andriole DA, Wathington HD, Tai RH. Educational outcomes for MD-PhD program matriculants: a national cohort study. Acad Med 2014; 89: 84–93.

Kwan JM, Noch E, Qiu Y, et al. The impact of COVID-19 on physician-scientist trainees and faculty in the United States: a national survey. Acad Med 2022; 97: 1536–45.

Milewicz DM, Lorenz RG, Dermody TS, Brass LF. Rescuing the physician-scientist workforce: the time for action is now. J Clin Invest 2015; 125: 3742–47.

Nguyen M, Cavazos JE, Venkataraman S, Fancher TL, Chaudhry SI, Desai MM, et al. Socioeconomic diversity in admissions to MD-PhD programs, 2014–19. JAMA Netw Open 2024; 7: e241951.

Pepin ME, Kamal Y, Reisman BJ, Rockman ME, Waller JP. Making the match or breaking it? Values, perceptions, and obstacles of trainees applying into physician-scientist training programs. medRxiv 2022; published online Aug 30. DOI:10.1101/2022.08.27.22278743 (preprint).

Sawalmeh M, Hatamleh H, Sawahreh H. Highlighting the need for MD-PhD programs in developing countries. Avicenna J Med 2023; 13: 65–67.

Unwin RJ, Challis B, MacPhee I, Hansen PBL, Jones R, Salama A, Barratt J. Enhancing collaboration between academia and industry in kidney disease research. Nephrol Dial Transplant 2025; 40: 836-838.

Unwin RJ, Challis B, Wagner C, Capasso G, Carlsson S. First-in-human to proof-of-concept: why experimental medicine studies remain essential in human physiology and drug development. Pflügers Archiv Eur J Physiol 2026; 478: 53.

Weggemans MM, Wolters FJ, Benedictus R, Prakken B, Ten Cate O, Pennings HJM. Motivation and determinants of research careers among physicians: exploring pros and cons of the Dutch MD-PhD model. PLOS One 2026; 21: e0343064.
